# Beyond the Scribe: Using Nominal Group Technique to Co-Design an Evaluation Framework for Patient-Held AI

**DOI:** 10.64898/2026.10.15.26365669

**Authors:** Catherine M. DesRoches, Amanda Norris, Jackie M. Barr, Will Vaughan, Roma Ruparel, Yun Liu, Matthew Thompson, Jennifer Stevens

## Abstract

The explosive growth of ambient AI scribes in healthcare has focused almost exclusively on relieving clinician documentation burdens and enhancing clinician efficiency and reducing burnout. While most research has focused on the clinician experience, these same types of tools, such as a patient-held AI Personal Health Assistant, can be put in the hands of patients to facilitate communication with healthcare providers, to provide greater autonomy and activation for patients in managing their own healthcare, and to facilitate the task of receiving and retaining complex new information. However, to ensure these tools serve as more than just administrative aids, we partnered with a large technology company to conceptually co-design how such a tool might be studied in real-world settings in a way that reflected outcomes of greatest importance to patients and families. We used the Nominal Group Technique NGT and engaged 33 patients and caregivers and 10 multidisciplinary clinicians in structured virtual sessions to identify the outcomes on which subsequent studies should hinge.

Our findings reveal a significant misalignment between current industry trends and patient priorities. While the market emphasizes the scribe function for billing, stakeholders overwhelmingly prioritized the AI-PHA’s role in longitudinal care management and post-visit coordination. The most critical outcome identified was the impact on communication and respect, suggesting that patients view AI as a mechanism to improve the human-to-human dynamic and ensure a shared understanding of the care plan. Our findings further highlighted that the primary hurdle for the use of patient-held AI is not technical utility but clinician trust and the need for formal consent processes.

Importantly, as this technology is new and integration into the clinical experience is still being explored, patients cannot assume they will have access to the transcript of their own encounter when that record is created and held by the health system. This gap highlights the potential value of a patient-held tool, preserving discussions from their visit and follow-up steps in clearer detail than they may receive in their clinical note or traditional visit summary.

From a system perspective, this transition requires a strategic shift from measuring workflow-focused efficiency to measuring patient-side advocacy and engagement. By centering the evaluation of AI on these patient-defined metrics and fundamentally designing subsequent AI investigations using a partnership model, health systems can move beyond simple automation toward a model of care delivery that empowers patients to navigate complex clinical journeys with greater autonomy and clarity.

## Background

Since the introduction of generative artificial intelligence (AI), its use in healthcare has grown exponentially, with the healthcare industry among the fastest adopters of such tools.^1^ Tools have been designed and developed for tasks across the healthcare spectrum, including making diagnoses, interpreting test results, relieving documentation burdens, and responding to patient messages.^2–7^ One area of particularly rapid growth is the use of AI-powered ambient scribes.^8^ These tools “listen” to the conversation between a patient and a clinician and produce a clinical note suitable for documentation and billing requirements. Research suggests that the use of an ambient scribe reduces clinician burnout and may reduce time spent in documentation. Click or tap here to enter text.^8–12^

The insertion of this third party into the clinician-patient relationship raises the question: how can ambient scribes help not just clinicians but also assist patients in engaging with and navigating our complex healthcare system? More than a decade of research has demonstrated that patients derive significant value from reading their clinical notes, including feeling more in control of their care, better prepared for visits, and more able to take their medications as prescribed.^13–17^ While patients with lower levels of education were just as likely, or in some cases more likely, to report benefits, concerns remain about the extent to which patients can understand and use notes containing complicated language and medical jargon, particularly for patients with limited English proficiency or lower levels of literacy.^18,19^ Generative AI and ambient scribes may offer a solution to this potential understanding gap. Just as large language models can be trained to produce a clinical note from a conversation, they could be trained to produce a patient visit summary. This summary, written and formatted for easy understanding, could contain clinically relevant information and next steps and help to improve patient activation through increased understanding.

### Co-designing an evaluation strategy with patients: an approach to measure patient-facing AI scribes

Research on the use of AI in healthcare has focused primarily on clinicians’ views and experiences, with little empirical work examining what patients want from such tools or how they would use them.^20^ For example, evaluations of the use of LLMs to create clinical notes and imaging reports for patients often include physician ratings of “readability” or “friendliness” but do not incorporate the perspectives of patients and their families, the intended beneficiaries of these tools. Click or tap here to enter text.^21–23^ This lack of inclusion may result in tools and products that do not meet the diverse needs of patients and families. Community engagement and co-design methods offer structured frameworks for incorporating the views and experiences of all stakeholders in processes such as product design, the prioritization of research questions, and the identification of outcomes important to the community.^24^ Structured consensus methods, such as the nominal group technique (NGT), provide a useful engagement approach for ensuring that diverse perspectives are systematically heard and integrated.^25^ Using this inclusive approach, done formally under an IRB waiver exemption, we identified what outcomes would be of greatest importance to a group of patients and clinicians when considering integrating a patient-facing AI scribe into care

### The Tool: AI Personal Health Assistant

In partnership with a large technology company, we sought to study the concept of an AI Personal Assistant (AI-PHA). This prototype application would be held on a patient’s mobile device and be designed for patient use before, during, and after clinical visits. AI-PHA would include an LLM-powered chatbot and an ambient scribe function. A patient would be able to use the chatbot to interact with AI-PHA before a clinical visit to prepare questions and review symptoms. During the visit, an ambient scribe function could record the conversation, track information and questions, and generate a summary. After the visit, patients could review the conversation, summary, and next steps, and use the chatbot to ask questions and set reminders for follow-up steps.

### Using NGT to codesign future patient-centered evaluations

Nominal group technique (NGT) is a structured group discussion used to generate ideas and reach consensus on their prioritization.^25^ In NGT groups, participants are asked to individually generate as many ideas as possible about a given topic in a specific amount of time. Each participant then reports their ideas to the group. The group then conducts at least two rounds of voting to reach a prioritized list. Time is allotted before each round of voting for members to ask questions and clarify ideas.

Participants in the stakeholder engagement activities were recruited from the Open Notes Labs Insight Panel. The Insight Panel is a group of patients, care partners, and practicing clinicians from around the United States who have agreed to be contacted to participate in stakeholder engagement and co-design activities. The project team sent the project plan and details to 70 Insight Panel members, and 43 opted into participation. Patient participants were grouped based on availability. All clinicians participated in one group.

Stakeholders participated in our groups virtually. The 90-minute groups were held in January 2026 and included the following compenents. First, participants viewed mockups of the use of the tool prior to, during, and after a clinical visit. (Figures 1 & 2) Second, participants watched a scripted clinical visit in which the patient introduced the concept of AI-PHA to the clinician and demonstrated how it could be used. Third, participants were asked to silently and individually generate as many ideas as possible within 2 minutes in response to the prompt: “How would this app help you in your healthcare experiences?” for patients, and “How would this app help patients?” for the clinician group. Each participant then read their ideas aloud, and each unique idea was posted to a virtual whiteboard by a member of the project team. Fourth, participants voted by selecting their top 3 to 4 ideas. Fifth, from this narrowed list, they selected their top 2 to 3 ideas, producing the final set of priorities. We then repeated this process with a second question: “If we were to study this tool, what outcomes would matter most to you—how would you know it’s helping (you/patients)?”

**Figure 1:**
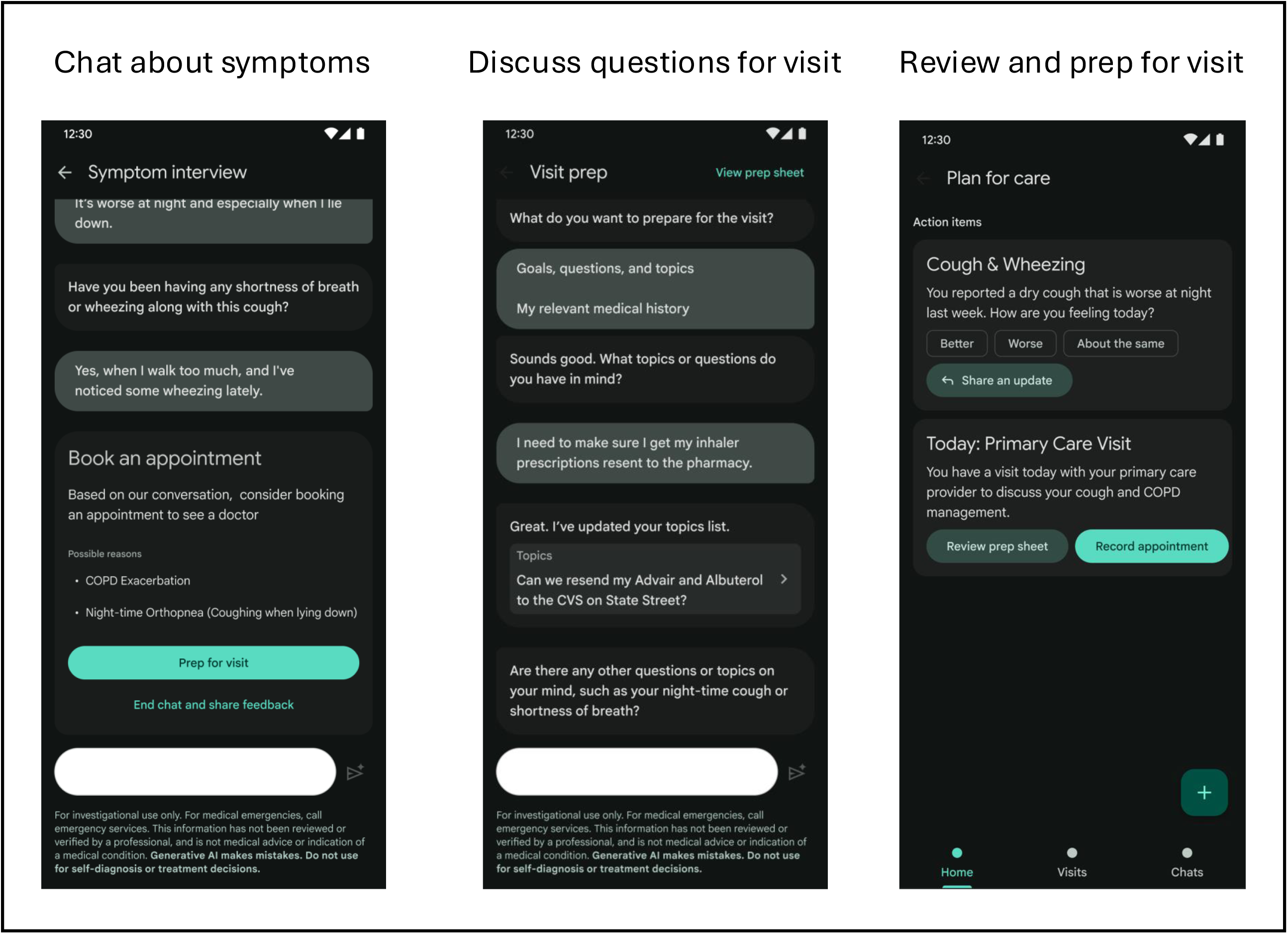
Prototype AI-PHA pre visit use.

**Figure 2:**
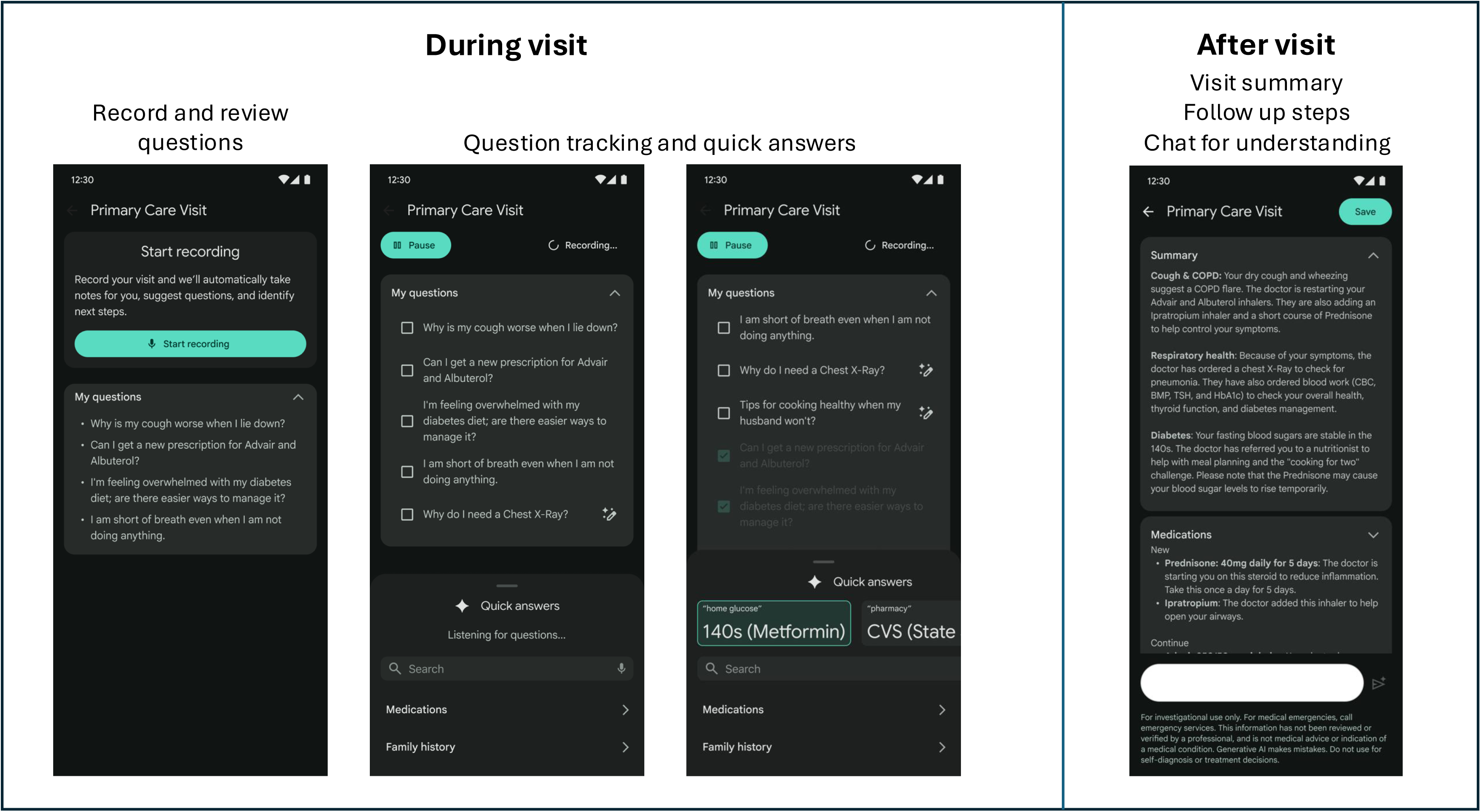
Prototype AI-PHA use during and after visits.

We analyzed our results in three steps. First, we reviewed all prioritized lists across the four groups. Three members of the research team independently grouped list items into themes and then met to discuss and reconcile their groupings, reaching consensus on a final thematic structure. Second, these themes served as a codebook for a structured review of the group transcripts, conducted by the same three team members, to identify any themes that emerged in discussion but were not reflected in the prioritized lists. A theme identified from the transcripts was added to the final set of priorities if it appeared in at least three of the four groups. The team met and resolved differences in coding through discussion. Outcomes were further grouped by type.

Thirty-three patients and 10 clinicians participated in their respective groups. Characteristics of the participants are shown in Table 1. The clinician group was comprised of a nurse, an informaticist, 5 physicians, and 3 mental health professionals. As shown in Table 1, participants were, on average, 45 years of age or older, female, spoke English at home, and had at least a college degree. The groups generated between 28 and 50 unique ideas for prompt 1 and 20 to 36 unique ideas for question 2 (Technical Appendix A). Through the voting process, the final number of prioritized ideas per group ranged from 7 to 15 for prompt 1 and 9 to 16 for prompt 2.

**Table 1:** Participant Characteristics.

| Characteristic |  | N |
| --- | --- | --- |
| Age | 18-34 | 6 |
|  | 35-44 | 6 |
|  | 45-54 | 12 |
|  | 55-64 | 10 |
|  | 65+ | 9 |
| Gender | Male | 10 |
|  | Female | 31 |
|  | Non-Binary or transgender | 2 |
| Primary language is English | Yes | 42 |
|  | No | 1 |
| Race | White | 19 |
|  | African American or Black | 10 |
|  | Asian | 5 |
|  | Hispanic | 2 |
|  | Other | 7 |
| Patient education | Highschool or less | 1 |
|  | Some college | 7 |
|  | Bachelor's Degree | 8 |
|  | Master's degree or more | 15 |
|  | Other | 2 |
| Clinician education | Master's Degree | 2 |
|  | Doctoral Degree | 8 |
|  | Other | 0 |
| Role in group | Clinician | 11 |
|  | Patient | 38 |
|  | Caregiver | 14 |
| Region | Northeast | 6 |
|  | Midwest | 11 |
|  | South | 10 |
|  | West | 16 |

### How can AI-PHA help patients?

Participants’ prioritized ideas were grouped into seven themes describing how AI-PHA could be useful to patients, ordered here from most to least number of ideas generated within the theme (Table 2). General care management was the most frequently prioritized theme, reflecting participants’ discussion of AI-PHA’s potential usefulness as a longitudinal tool extending beyond the clinical visit. Patients described the potential to use the app to maintain their own records of past treatments and medications, track symptoms and health trends over time, and support medication reconciliation. Several participants emphasized the value of having a single, reliable record to bring to different providers. As one patient noted, ’I sure don’t remember the number of visits I’ve had over the years with this condition, so I’d like a historical record in one place.’ Patients also described wanting to use the tool to communicate more effectively with specialists and other members of their care team, and to advocate for themselves when navigating difficult conversations. One patient captured this directly: ’I wouldn’t have to battle so much for medications that I need, or procedures, because I would be able to convey this to people who don’t understand my condition.’ Shareability was a consistently upvoted feature in this theme, with patients noting they would use AI-PHA to share information with friends and family involved in their care.

**Table 2:** Prioritized uses for AI-PHA and illustrative quotes, sorted in descending order by the frequency at which ideas within each theme were surfaced.

| Theme | Identified by |  | Illustrative quote |
| --- | --- | --- | --- |
|  | Clinicians | Patients |  |
| Visit preparation | x | x | It would help me prepare for my visit better with even more personalized questions about what I want to know for my health. [patient]<br>I would use it to triage the symptoms I have. [patient]<br>Help patients to prepare and remember questions [clinician] |
| Visit support | x | x | Seeing that you can make a list and record in the same app is great, and that the app can help check things off your list. [patient]<br>It helps the patients ask better questions. That means they will be more confident during the appointment. [clinician] |
| Post visit care coordination | x | x | I see it helping you talk with other health professionals like your nutritionist or being able to talk to your pharmacist about things that were brought up in the visit that you didn't remember. [patient]<br>Being able to share it with friends and family because they always say, "Did you ask this? Did you forget to ask that?" There's always a million questions from your family. [patient]<br>Telling them who to follow up with and why, which patients might not always remember [clinician] |
| General care management |  | x | I wouldn't have to battle so much for, let's say, medications that I need, or procedures because I would be able to convey [this] to people who don't understand my condition or believe that I have it. [patient]<br>I sure don't remember the number of visits I've had over the years with this [condition], so I'd like a historical record in one place. [patient] |
| Business of healthcare |  | x | And just having a running record of the finances because you know what tests were done, what tests were talked about, all of that. [patient]<br>I like it has been recorded basically by a third party, and not just going back to a visit note, where the doctor may not have transcribed something that happened during the visit. [patient] |
| Accessibility and usability | x | x | I'm physically handicapped, so [ . . . ] to write things down when I'm alone is difficult. Having something record visit instructions and everything is very handy. [patient]<br>It would be very useful for people with cognitive impairments, autism, mental illness, etc, where incoming information is hard to digest. [clinician] |
| Accuracy |  | x | This is a little more true to what actually occurred. [patient]<br>I think it's important if the doctor mentions those modalities [additional / alternative therapies] to actually capture that in the summary because I think those discussions happen, but it doesn't get into the medical record the same way. [patient] |

Post-visit care coordination was the second-most-frequently prioritized theme. Participants noted that they would use AI-PHA to remember what happened during the visit, understand the rationale for their treatment plan, and track follow-up tasks and referrals. The tool’s potential to help patients navigate the period immediately after a visit was a recurring discussion point. One patient described the possibility of using the app to talk with other healthcare professionals after a visit: ’I see it helping you talk with other health professionals like your nutritionist, or being able to talk to your pharmacist about things that were brought up in the visit that you didn’t remember.’ Participants also noted that AI-PHA could increase referral follow-through by helping patients prioritize next steps and understand who to contact with questions.

Accessibility was the third-most-prioritized theme. Participants identified that the tool could be particularly valuable for patients with physical disabilities, cognitive impairments, or conditions such as autism or mental illness that make it difficult to process information in real time. One patient with a physical disability noted that having something to record visit instructions would be ’very handy’ given the difficulty of writing things down when alone. Clinicians echoed these priorities, noting the tool’s potential for patients with cognitive impairments and those facing language barriers in clinical settings.

While respondents understood that uses such as post-visit coordination required the tool’s record function to be turned on, otherwise engaging with the tool directly during the visit was the fourth-most-prioritized theme. Participants noted that they would use AI-PHA to track the conversation in real time, ask better questions, and feel more confident during the visit. Several noted the value of being able to check questions off a list as they were addressed. One patient described the appeal of having the list and recording function in a single app: ’Seeing that you can make a list and record in the same app is great, and that the app can help check things off your list.’ Clinicians similarly noted that the tool could help patients feel more confident and ask better questions during appointments.

The theme of pre-visit preparation was prioritized less frequently but was identified by both patients and clinicians. Participants valued the possibility of using AI-PHA to prepare personalized questions for their visit, triage symptoms, and think through whether and where to seek care. Clinicians noted the value of patients arriving better prepared, with relevant questions already organized.

Accuracy of the AI-PHA’s transcription and summarization features was the least frequently prioritized theme, although it was discussed in all groups. Patients discussed accuracy in the context of the business of healthcare reflecting concerns about accountability, record accuracy, and the financial dimensions of care. Patients were positive about the potential utility of the tool in maintaining accurate records through documenting what actually occurred during a visit, particularly when the clinical note might not fully reflect the conversation, noting that this could be useful for insurance claims and denials. Further, several patients noted specifically that the app would capture the clinician’s rationale for treatment recommendations and any discussion of alternative or complementary therapies, things that are not typically included in a clinical note. As one patient observed, the app could be “a little more true to what actually occurred.”

### What outcomes are most important?

The outcomes participants prioritized reflected both practical and relational dimensions of care, cutting across traditional measurement categories such as patient-reported outcomes, healthcare process measures, and service utilization.

The most highly prioritized outcome theme across all four groups was communication; specifically, whether AI-PHA could help improve the quality of understanding and communication between patients and clinicians (Table 3). Participants described wanting to leave visits with a clear, shared understanding of their care plan. As one patient noted, the value of the tool was in ’leaving settled, feeling heard, with a clear path forward.’ Patients also emphasized the relational dimension of this outcome, with several noting that the tool had the potential to improve the dynamic between patient and clinician, shifting what one participant described as a ’fight or push and pull’ toward a more collaborative exchange. Language accessibility was also a recurring concern within this theme, with participants noting that the tool could help bridge the gap between medical terminology and plain language, particularly for patients for whom English is a second language. One patient described how the recording could allow them to ’go back to understand’ what was said, and that this understanding in turn would allow them to ‘make better’ decisions about their care.

**Table 3:** Prioritized outcome themes with illustrative quotes, by stakeholder group.

| Theme | Measurement Category | Identified by Patients | Identified by Clinicians | Illustrative Quotes |
| --- | --- | --- | --- | --- |
| **Communication** | *Patient-reported outcomes* | ✓ | ✓ | "Leaving 'settled,' feeling heard, with a clear path forward." [patient]<br>"...improves the dynamic between patient and doctor, so it's not a fight or push and pull." [patient] |
| **Usability and engagement** | *Accessibility and usability* | ✓ | ✓ | "Do I still want to have this app after I've used it for a month." [patient]<br>"But would feel useful in the long term and not one more thing to remember." [patient] |
| **Outcomes and utilization** | *Healthcare utilization; clinical outcomes* | ✓ | ✓ | "Over the course of managing a disease, one of the key metrics I look for is fewer ER visits or hospitalizations." [patient]<br>"My condition would be improved. I might be able to resume my lifestyle." [patient] |
| **Care coordination** | *Healthcare process measures* | ✓ | ✓ | "For me, it's being able to communicate with my other doctors and know that I'm not guessing what the first one said." [patient]<br>"Can the patient explain the next steps to the nurse or team." [clinician] |
| **Patient burden** | *Patient-reported outcomes* | ✓ |  | "I know it would be helpful if it helped with my medical burnout as a chronic illness patient." [patient]<br>"Less time, less stress, less time in the communication side of diagnostics." [patient] |
| **Adherence and actionable plan** | *Healthcare process measures* | ✓ | ✓ | "Does the patient make better choices about what to do next." [patient]<br>"Adherence to next steps — get new medication, follow up on referral, etc." [clinician] |
| **Workflow and provider experience** | *Provider-level outcomes* | ✓ | ✓ | "I think this would help move that along for patients who don't really know what they want to say." [patient]<br>"Reduce the number of calls, emails, and repetitive questions." [clinician] |

*Note: Measurement categories correspond to the broader outcome themes shown in Table 5. Some granular themes map onto more than one measurement category.*

Usability, including potential ongoing engagement with AI-PHA, was the second most highly prioritized theme. Key outcomes included whether patients would continue to use the tool after their initial visit, whether they felt more confident and in control of their healthcare, and whether the theoretical tool felt genuinely useful rather than burdensome. One patient captured this tension directly, noting that a meaningful outcome would be whether the tool felt ’useful in the long term and not one more thing to remember.’ Clinicians echoed these priorities, emphasizing patient satisfaction, ease of use for older patients, and whether the tool’s output was clear, concise, and free from jargon.

Health outcomes and service utilization was the third most prioritized theme and spanned both patient-reported and healthcare utilization measures. Patients identified a range of outcomes, from concrete measures of healthcare utilization (e.g., fewer emergency department visits, fewer hospitalizations, fewer missed appointments) to functional improvements (e.g., return to work or activity) and disease-specific improvements (e.g., blood pressure or blood sugar at target). Several patients framed these potential outcomes in terms of what mattered most to their daily lives. One participant stated: ’What matters to me is that my condition would be improved. I might be able to resume my lifestyle.’ Another identified ’better sleep, more energy, weight loss’ as the outcomes they would use to know the tool was helping. Clinicians similarly prioritized utilization outcomes, focusing on reductions in unnecessary return visits and specialty referrals.

Care coordination outcomes were prioritized by both patients and clinicians, though with differing emphases. Patients focused on the tool’s potential to help them communicate across providers by sharing accurate information with specialists, pharmacists, and other members of their care team, as well as on preserving information across insurers and avoiding duplicative testing. As one patient described it, the tool would allow them to ’close the communication loop’ between physicians. Clinicians focused on whether patients could accurately explain their next steps to the care team and whether the tool reduced unnecessary follow-up contacts, such as portal messages and phone calls.

Patient burden was consistently prioritized across all three patient groups. Participants described outcomes including reduced patient stress, time savings, and relief from the cognitive and administrative demands of navigating the healthcare system. One patient with a chronic illness described hoping the tool would help with ’medical burnout.’ Others pointed to the practical burdens of managing multiple portals and providers, with one participant noting that a meaningful outcome would be ‘transferring information’ seamlessly. The care partner dimension of burden also emerged, with participants noting that the tool could reduce stress not only for patients but for family members involved in their care.

Adherence and actionable plan outcomes were prioritized by both groups, centering on whether patients left visits with a clear, achievable plan and followed through on it. Specific outcomes included completed referrals, filled prescriptions, and attendance at follow-up appointments. Clinicians emphasized measurable adherence indicators such as completion of next steps. Patients framed these outcomes more personally, focusing on whether the tool helped them ’make better choices about what to do next’ and remember what they were supposed to do after the visit.

Workflow and provider experience was identified by both patients and clinicians, though it ranked lower for both. Outcomes in this theme included visit length, inbox message volume, repetitive questions, and visit efficiency. The relatively low prioritization of workflow outcomes suggests that both patients and clinicians viewed the primary value of AI-PHA as potentially improving the patient experience rather than streamlining clinical operations.

### The role of physician consent

Transcript review revealed an additional theme that was not included in the prioritized list. Across all three patient groups, participants raised the issue of physician consent to be recorded as a critical factor to assess. While participants viewed AI-PHA as a potentially useful tool for managing their care, they noted that its utility would be diminished if physicians did not consent to being recorded.

### From priorities to a phased research approach

The outcome themes identified by participants were further grouped into broader measurement categories to facilitate the development of a research roadmap, as the study design required varies considerably across outcome types (Table 4). The breadth of outcomes prioritized by patients and clinicians reflects the complexity of evaluating a tool like AI-PHA and suggests that a phased research approach will be necessary. Some primary outcomes, such as tool usability, and summary accuracy, are appropriate for an early feasibility study and can be assessed without a comparison group or a large sample size. Others, such as medication adherence, patient-reported quality of life, and patient activation, require a randomized study design to attribute change to the tool. Still others, such as hospital readmission rates and disease control, are relatively rare events or require longer follow-up periods and larger sample sizes, making them more appropriate for a fully powered multi-site clinical trial. Table 4 maps the prioritized outcome themes onto a three-phase research program, from a single-site feasibility study through a pilot randomized controlled trial to a definitive multi-site clinical trial.

**Table 4:** NGT-prioritized outcome themes mapped to a phased research program, by stakeholder group.

| Outcome Theme | Example Outcomes | Identified By | Phase 1<br>*Single-site feasibility*<br>*No comparison group;*<br>*process outcomes* | Phase 2<br>*Pilot RCT*<br>*Randomized;*<br>*preliminary efficacy* | Phase 3<br>*Multi-site RCT*<br>*Powered for clinical and*<br>*utilization outcomes* |
| --- | --- | --- | --- | --- | --- |
| **Safety and accuracy** | Summary is error-free; summary reflects patient's primary concerns | *Patients and clinicians* | ✓ (priority) | ✓ | ✓ |
| **Overall tool utility** | Percent of visits in which tool is used; tool use within 7 days pre and post visit | *Patients only* | ✓ | ✓ (priority) | ✓ (priority) |
| **Patient-reported outcomes** | Change in satisfaction, patient activation, stress, and quality of life | *Patients and clinicians; quality of life identified by patients only* |  | ✓ (priority) | ✓ (priority) |
| **Provider-level outcomes** | Visit length; inbox message volume | *Patients and clinicians* |  | ✓ | ✓ (priority) |
| **Healthcare process measures** | Medication adherence; follow-up completion rate | *Patients and clinicians* |  | ✓ | ✓ (priority) |
| **Care partner and family outcomes** | Summary shared with care partner | *Patients only* |  | ✓ | ✓ |
| **Clinical outcomes** | Condition controlled or at target; functional improvement | *Patients only* |  |  | ✓ (priority) |
| **Healthcare utilization** | Hospital readmission rates; volume of return visits | *Patients and clinicians* |  |  | ✓ |
| **Physician consent to recording†** | Clinician consent rate | *Patients only (transcript review)* | ✓ | ✓ | ✓ |

†Physician consent to recording was identified through transcript review as a design consideration rather than a study outcome, and informed the recruitment strategy for Phase 1.

### The urgency to engage patients in co-designing the evaluation of AI tools

We describe a structured stakeholder engagement process to generate and prioritize use cases and outcomes for evaluating a patient-held AI health companion, demonstrating that it is both feasible and meaningful to include patients and caregivers as active partners in the design of research intended to benefit them. Artificial intelligence seems poised to transform every facet of health care, and the speed with which AI-powered tools are being adopted across the industry is notable.^1^ Given the speed of this transformation, we may wonder whether careful and rigorous evaluation of new tools is still relevant. If we are living in the era in which “moving fast” is over-emphasized by some, is the time required to thoughtfully examine how such tools affect patients and clinicians still available and necessary? Results from this co-design exercise and others suggest that the answer to this question should be yes.^26,27^

Findings from this work echo and extend public polling on the use of AI in healthcare, indicating that patients are interested in and willing to use AI to help understand and engage in their healthcare.^28–30^ However, they express caution about trusting AI-powered tools and expect them to be accurate.^31–33^ Assessing accuracy and usability requires careful research methods and independent investigations. Without them, patients are left to navigate a “buyer beware” environment in which they must evaluate accuracy and utility, a task that would be challenging for most.

Uncertainty around the effects of AI powered tools in health care extends beyond accuracy to questions of access and control. Since this technology is new and integration into the clinical experience is still being explored, patients cannot assume they will have access to the transcript of their own encounter when that record is created and held by the health system.^34^ This gap highlights the potential value of a patient-held tool, preserving discussions from their visit and follow-up steps in clearer detail than they may receive in their clinical note or traditional visit summary.

The use of NGT to inform study design has limitations. First, NGT produces a list of ideas within each group, allowing us to describe each group’s priorities. However, the process does not allow for statistical comparison of priorities across groups. Second, the structure of the voting process allows the groups to reach consensus but may also result in the loss of relevant and important ideas that did not rise to the top in voting. Our transcript review was designed in part to address this limitation. Third, the virtual administration of the groups may have affected group dynamics as compared to in-person meetings. Fourth, while our patient participants were diverse in their experience with the healthcare system, they were not a representative sample. Individuals willing to participate in a co-design process may hold different perspectives than those who are not and may differ in ways that are not measurable. Similarly, while our clinician group was multidisciplinary, it was a single group and may not reflect the full range of their perspectives. Finally, our ratio of patient to clinician groups may have led to a de-emphasis of themes and outcomes important to clinicians.

The findings reported here are themselves a contribution to the field. Researchers designing evaluations of AI-powered tools intended for patients can draw on the themes and specific outcomes identified as a framework for design. More broadly, our work demonstrates that it is both feasible and valuable to include patients and caregivers as active partners in the design of research intended to benefit them. In an era defined by the rapid adoption of AI in healthcare, the imperative to evaluate these tools carefully, and to do so in partnership with the patients they are meant to serve, has never been greater.^35^ Moving fast may be inevitable; but it cannot be at the expense of breaking patients’ trust, safety, and care. Instead, we should strive to move at the speed of trust and evidence, using the diverse needs of both patients and clinicians as a guide.

Technical Appendix A

## Data Availability

Data produced in the present study are not available due to privacy restrictions placed on qualitative data.

## Appendix A: Nominal group technique idea prioritization

Technical Appendix A Appendix A: Nominal group technique idea prioritization
|  | How would this app help you/patients? |  |  | What outcomes would matter most to you? |  |  |
| --- | --- | --- | --- | --- | --- | --- |
|  | Number of ideas generated | 1<sup>st</sup> Vote | 2<sup>nd</sup> Vote | Number of ideas generated | 1<sup>st</sup> Vote | 2<sup>nd</sup> Vote |
| Patient group 1 | 50 | 30 | 15 | 35 | 18 | 11 |
| Patient group 2 | 30 | 22 | 11 | 25 | 21 | 12 |
| Patient group 3 | 31 | 26 | 14 | 29 | 23 | 16 |
| Clinician group | 28 | 23 | 7 | 20 | 15 | 9 |

